# Clinical features and sexual transmission potential of SARS-CoV-2 infected female patients: a descriptive study in Wuhan, China

**DOI:** 10.1101/2020.02.26.20028225

**Authors:** Pengfei Cui, Zhe Chen, Tian Wang, Jun Dai, Jinjin Zhang, Ting Ding, Jingjing Jiang, Jia Liu, Cong Zhang, Wanying Shan, Sheng Wang, Yueguang Rong, Jiang Chang, Xiaoping Miao, Xiangyi Ma, Shixuan Wang

## Abstract

**Background:** As of March 2, 2020, SARS-CoV-2 has infected more than 80174 people and caused 2915 deaths in China. This virus rapidly spreads to 56 countries worldwide. Thus, in order to effectively block its transmission, it is urgent to uncover all the possible transmission routes of SARS-CoV-2.

**Methods:** From January 28 to February 18, 2020, 35 female patients diagnosed with COVID-19 in Tongji Hospital were included in this descriptive study. The gynecologic history, clinical characteristics, laboratory findings and chest computed tomography (CT) of all patients were recorded in detail. To examine whether there is sexual transmission through vaginal from female to her partner, we employed real-time polymerase chain reaction testing (RT-PCR) to detect SARS-CoV-2 in vaginal environment (including vaginal discharge, cervical or vaginal residual exfoliated cells) and anal swab samples, and inquired recent sexual behaviors from the patients.

**Findings:** The age range of the 35 patients with COVID-19 was 37-88 years. Over 50% patients infected with SARS-CoV-2 had chronic diseases. We tested the vaginal environment and anal swabs from the 35 female patients with COVID-19 and found that only an anal swab sample from one patient was positive for SARS-CoV-2. All the samples from vaginal environment were negative for SARS-CoV-2. The infection rate of the patients’ sexual partner was 42·9%. Additionally, two female patients admitted having sex with their partners during a possible infection incubation period, while one patient’s partner was uninfected and the other patient’s partner was diagnosed with COVID-19 (after the diagnosis of the female patient).

**Conclusion:** No positive RT-PCR result was found in the vaginal environment perhaps due to the lack of ACE2 expression, which is the receptor of SARS-CoV-2, in the vagina and cervix tissues (human protein atlas). The results from this study show no evidence of transmission of SARS-CoV-2 through vaginal sex from female to her partner. However, the risk of infection of non vaginal sex and other intimate contacts during vaginal sex should not be ignored.

**Funding:** This work was financially supported by the Clinical Research Pilot Project of Tongji hospital, Huazhong University of Science and Technology (No. 2019CR205).

## Introduction

A novel coronavirus (severe acute respiratory syndrome coronavirus 2, SARS-CoV-2) infectious disease has erupted in Wuhan, Hubei Province since December 2019^1,2^. It has spread rapidly throughout entire China. Up to 20:00 on March 2, 2020, the total number of confirmed cases of COVID-19 in China has reached 80174 with 2915 deaths ^3^. Among the infected proportion, 48.6% are females ^4^. Internationally, cases of COVID-19 have been reported in 56 countries, especially in Japan, South Korea and Singapore, with rapidly increasing trend. SARS-CoV-2 is so widely spread, leading to injury of multiple organs and deaths. Therefore, it is crucially important to identify the transmission routes of SARS-CoV-2 in order to prevent its further spreading.

Previously in 2003, there was an outbreak of SARS-CoV in China ^5^. The main transmission route of SARS-CoV was via respiratory droplets, although the potential of transmission by opportunistic airborne routes and environmental factors was also reported ^6,7^. In addition, SARS-CoV was detected in urine, feces and tears of some SARS-CoV infected patients. It was confirmed that SARS-CoV could be transmitted through fecal-oral and contiguous pathways ^6,8^. Thus far, there has not been a report to show if SARS-CoV can be transmitted through sexual transmission. Therefore, based on the transmission route of SARS-CoV, current preventive measures, including maintaining good personal and environmental health, and implementing strict contact and droplet prevention measures in the community, can effectively prevent the spread of SARS-CoV^6^. However, different from SARS-CoV, MERS-CoV, another virus of the same coronavirus family, which was first reported in Saudi Arabia in 2012 and later spread to several other countries^9,10^, was transmitted through large droplets and close contact, although the possibility of air transmission or dust mite transmission was not ruled out^9^. Also, it is not clear whether MERS-CoV is sexually transmitted. The World Health Organization (WHO) has provided recommendations for the prevention of MERS-CoV, mainly including droplet and contact precautions^11^. Interestingly, HCoV-229E in the coronavirus family was detected in the maternal vagina and also found in the gastric swab samples of newborns ^12^. This makes the sexual transmission of this type of virus possible although there is no direct evidence to prove it. Therefore, it also remains possible that SARS-CoV-2, as a new type of coronavirus, could be sexually transmitted.

Currently, the main known transmission route of SARS-CoV-2 is through respiratory air droplets. The other routes could be through secretary liquids, such as from eyes^13,14^. SARS-CoV-2 was shown to exist in the feces of patients with COVID-19, suggesting that fecal-oral transmission of SARS-CoV-2 is possible ^15^. Although there is currently no evidence for vertical transmission ^16^, it still remains unknown whether SARS-CoV-2 has other transmission routes from person to person. To explore the possibility of sexual transmission of this contagious virus, we performed RT-PCR analysis of SARS-CoV-2 in the anal swab samples and vaginal environment samples (including cervical or vaginal residue exfoliated cells, vaginal discharge) collected from 35 female patients with COVID-19 and inquired their recent sexual behavior.

## Methods

### Study design and patients

In this study, we recruited 35 female COVID-19 patients from January 28 to February 18, 2020 in three branches of Tongji Hospital, Sino-French New City Branch (16 cases), Optical Valley Branch (16 cases) and Hankou main campus (3 cases) respectively, which are located in different geographical areas of Wuhan. Cases of COVID-19 were diagnosed based on the New Coronavirus Pneumonia Prevention and Control Program (5th edition) published by the National Health Commission of China^17^. Twenty-seven patients with COVID-19 were tested SARS-CoV-2 positive by RT-PCR analysis on samples from the respiratory tract, and eight patients were diagnosed as clinically confirmed cases based on the 5th guidance mentioned above according to epidemiological history, symptoms and chest CT. This study was reviewed and approved by the Medical Ethical Committee of Tongji Hospital of Huazhong University of Science and Technology (TJ-IRB20200208). The trial has been registered in Chinese Clinical Trial Registry (ChiCTR2000029981). Written informed consent was obtained from each enrolled patient.

### Data collection

The gynecologic history, clinical characteristics, laboratory findings, chest computed tomography (CT) and outcome data were obtained from patients’ records. To ascertain the authenticity and integrity of the gynecologic history, especially the sexual behavior history, we also directly communicated with patients or their families. All information was obtained and curated with a standardized data collection form. Two researchers also independently reviewed the data collection forms to double check the data.

Vaginal discharge samples from patients with COVID-19 were obtained from posterior fornix of vagina according to the protocol of the virus sampling kit (Yocon, Beijing, China). Exfoliated cell samples were collected from cervix or vaginal residue for patients undergoing hysterectomy according to the protocol of Thinprep cytologic test (Hologic, Massachusetts, USA)^18^. Throat swab and anal swab samples were collected respectively from the upper respiratory tract and perianal tissues following the protocol of the virus sampling kit mentioned above. Diagnosis of COVID-19 was based on SARS-CoV-2 test of throat swab. Vaginal discharge, exfoliated cell and anal swab samples were collected about one week since diagnosed. All samples were tested for SARS-CoV-2 with the Chinese Center for Disease Control and Prevention (CDC) recommended Kit (DAAN GENE, Guangzhou, China or BioPerfectus Technologies, Jiangsu, China), following WHO guidelines for real-time RT-PCR. RT-PCR testing was used to detect COVID-19 according to the recommended protocol. The samples were processed simultaneously at the Department of Clinical Laboratory of Tongji Hospital and Wuhan KDWS Biological Technology Co, Ltd. Sample collection, processing, and laboratory testing complied with WHO guidance. Positive confirmatory cases of COVID-19 infection were defined as those with a positive test result from either laboratory.

### Statistical analysis

SPSS software, version 20·0, was used for statistical analysis. The continuous measurements were presented as mean (SD) if they were normally distributed or median (IQR) if they were not, and categorical variables as count (%). For laboratory results, we also assessed whether the measurements were outside the normal range.

## Results

35 patients with COVID-19 were recruited in this study, the range of their age was 37-88, with a mean age of 61·5 years (SD =11·2; Table 1). Most patients were natives of Wuhan (n =29, 82.9%), and only one had directly contacted with Huanan seafood market. The most common chronic medical diseases were hypertension (n =12, 34.3%) and diabetes (n =7, 20%). As for gynecological diseases, uterine fibroids were mentioned the most (n =13, 37·1%). Other diseases included cervical intraepithelial neoplasia (CIN) III, endometrial atypical hyperplasia, and endometrial cancer, with one patient for each disease. Most patients had entered menopause (n =28, 80%). For the rest 7 patients, except for 1 puerperal woman who had a cesarean section 2 days before with breast milk and neonatal throat swab SARS-CoV-2 negative, the remaining 6 patients still had menstruation, in which half of them complained of decreased menstrual flow.

**Table 1:** Demographics, baseline characteristics, and clinical outcomes of 35 Patients

|  | Patients (n =35) |
| --- | --- |
| **Age, years** |  |
| Mean (SD) | 61·5 (11·2) |
| Range | 37-88 |
| <45 | 4 (11·4%) |
| 45-55 | 5 (14·2%) |
| >55 | 26 (74·3%) |
| **Occupation** |  |
| Retired personnel | 26 (74·3%) |
| Professional/technical staff | 4 (11·4%) |
| Agricultural worker | 2 (5·7%) |
| Business/service worker | 3 (8·6%) |
| **Wuhan native** | 29 (82·9%) |
| **Exposure to Huanan seafood market** | 1 (2·9%) |
| **Chronic medical illness** |  |
| Hypertension | 12 (34·3%) |
| Diabetes | 7 (20·0%) |
| Coronary heart disease | 3 (8·6%) |
| Lymphoma | 1 (2·9%) |
| With any illness above | 19 (54·3%) |
| **Gynecological disease** |  |
| Uterine fibroids | 13 (37·1%) |
| CIN III | 1 (2·9%) |
| Endometrial atypical hyperplasia | 1 (2·9%) |
| Endometrial cancer | 1 (2·9%) |
| None | 19 (54·3%) |
| **Menstruation** |  |
| Menopause | 28 (80%) |
| Increased menstrual flow | 0 (0%) |
| Decreased menstrual flow | 3 (8·6%) |
| Constant menstrual flow | 3 (8·6%) |
| Puerpera* | 1 (2·9%) |
| **Sexual partner infection** | 15 (42·9%) |
| **Admitted sex behavior** | 2 (5·7%) |
| **Clinical outcomes** |  |
| Remain in hospital | 34 (97·1%) |
| Discharged | 1 (2·9%) |
| Died | 0 (0%) |

Data are n (%) unless specified otherwise.
\***Breast milk and neonatal throat swab samples was tested negative for SARS-CoV-2.**
†Time window: from 14 days before first symptoms to the day we collected samples.

The most common first symptom was fever (n =25, 71·4%), Due to the special diagnostic principles in Wuhan; only 27 (77·1%) patients were tested positive for SARS-CoV-2 (Throat swab). The remaining 8 patients were clinically diagnosed depend on the images of CT scan (Table2). All patients in this study were classified at admission, including mild group (n =1, 2·9%), severe group (n =32, 91·4%) and critical group (n =2, 5·7%, Table 2). After admission, the white blood cell count was decreased in 5 (14·3%) patients and increased in 4 patients (11·4%, Table 3). 16 (45·7%) patients had a lymphocyte count reduction, while 18 patients (51.4%) had a lymphocyte ratio below the normal range. C-reactive protein (CRP) level was increased in 14 (40%) patients. The erythrocyte sedimentation rate accelerated in 19 (54·3%) patients. 10 (28.6%) patients had increased aspartate transaminase (AST), and 13 (37·1%) patients had elevated alanine transaminase (ALT). The serum potassium was decreased in 5 patients (14·3%) and increased in 3 patients (8·6%). Five (14·3%) patients had decreased serum sodium. 28 of the 35 patients were tested for multiple cytokines, including Interleukin1β (IL-1β), Interleukin2R (IL-2R), Interleukin6 (IL-6), Interleukin8 (IL-8), Interleukin 10 (IL-10) and Tumor necrosis factor α (TNF-α). IL-2R, IL-6, and TNF-α were higher than the normal range in half or more patients, respectively. Notably, 9 (32·1%) patients’ level of IL-2R, IL-6, and TNF-α increased altogether. Every patient showed typical findings of chest CT images-multiple patchy ground-glass shadows in lungs. Typical images from each group were demonstrated (Figure 1).

**Table 2:** Clinical characteristics and treatment

|  | Patients (n =35) |
| --- | --- |
| **First symptom** |  |
| Fever | 25 (71·4%) |
| Muscle ache | 4 (11·4%) |
| Cough | 4 (11·4%) |
| Fatigue | 1 (2·9%) |
| Shortness of breath | 1 (2·9%) |
| **SARS-COV-2 (Throat swab) positive cases** | 27 (77·1%) |
| **Clinically diagnosed cases** | 8 (22·9%) |
| **Clinical group** |  |
| Mild | 1 (2·9%) |
| Severe | 32 (91·4%) |
| Critical | 2 (5·7%) |
| **Treatment** |  |
| Antiviral treatment | 33 (94·3%) |
| Antibiotic treatment | 29 (82·9%) |
| Glucocorticoids | 13 (37·1%) |
| Intravenous immunoglobulin therapy | 9 (25·7%) |
| Anticoagulant therapy | 3 (8·6%) |
| **Oxygen therapy** |  |
| Face mask/nasal catheter | 33 (94·3%) |
| BiPAP | 2 (5·7%) |
| Invasive ventilation | 0 (0%) |

**Table 3:** Laboratory results

|  | Patients (n =35) |
| --- | --- |
| **ITEM (Unit, Normal range)** |  |
| **White blood cell count (×10<sup>9</sup>/L, 3·5-9·5)** | 6·4(3·1) |
| Decreased | 5(14·3%) |
| Increased | 4(11·4%) |
| **Lymphocyte count (×10<sup>9</sup>/L, 1·1-3·2)** | 1·1(0·7-1·9) |
| Decreased | 16(45·7%) |
| Increased | 3(8·6%) |
| **Lymphocyte ratio (% , 20·0-50·0)** | 19·7(8·9) |
| Decreased | 18(51·4%) |
| **C-reactive protein concentration (mg/L, <10)** | 33(2·0-64·9) |
| Increased | 14(40%) |
| **Erythrocyte sedimentation rate (mm/H, <20)** | 47·2(33·0) |
| Increased | 19(54·3%) |
| **Procalcitonin (ng/mL, <0·5)** | 0·06(0·04-0·09) |
| 0·5-2 | 2(5·7%) |
| ≥2 | 0(0%) |
| **Alanine aminotransferase (U/L; ≤33)** | 26(18-41) |
| Increased | 10(28·6%) |
| **Aspartate aminotransferase (U/L; ≤32)** | 25(13-37) |
| Increased | 13(37·1%) |
| **Creatinine (umol/L, 45-84)** | 58(53-68) |
| Increased | 1(2·9%) |
| **Blood potassium (mmol/L, 3·5-5·1)** | 3·9(3·7-4·3) |
| Decreased | 5(14·3%) |
| Increased | 3(8·6%) |
| **Blood sodium (mmol/L, 136-145)** | 138·8(137·1-140·8) |
| Decreased | 5(14·3%) |
|  | **Patients (n =28)** |
| **Interleukin1β (pg/ml, <50)** | <5 (<5 - <5) |
| Increased | 5 (17·9%) |
| **Interleukin2R (u/ml, 223-710)** | 695·0 (458·5 -1038·8) |
| Increased | 14 (50%) |
| **Interleukin6 (pg/ml), <7)** | 10·6 (2·5 -29·1) |
| Increased | 15 (53·6%) |
| **Interleukin8 (pg/ml, <62)** | 11·1 (5·9-21·8) |
| Increased | 0 (0%) |
| **Interleukin 10 (pg/ml, <9·1)** | 5·0 (5·0-7·2) |
| Increased | 2 (7·14%) |
| **Tumor necrosis factor α (pg/ml, <8·1)** | 8·1 (6·6-10·2) |
| Increased | 14 (50%) |
| **Interleukin2R, Interleukin6, and Tumor necrosis factor α increased** | 9 (32·1%) |

Data are n (%), n/N (%), mean (SD), and median (IQR). Decreased means below the lower limit of the normal range and increased means over the upper limit of the normal range. Number of cases were 35 in blood routine and biochemistry tests, 28 in cytokines tests.

**Figure 1:**
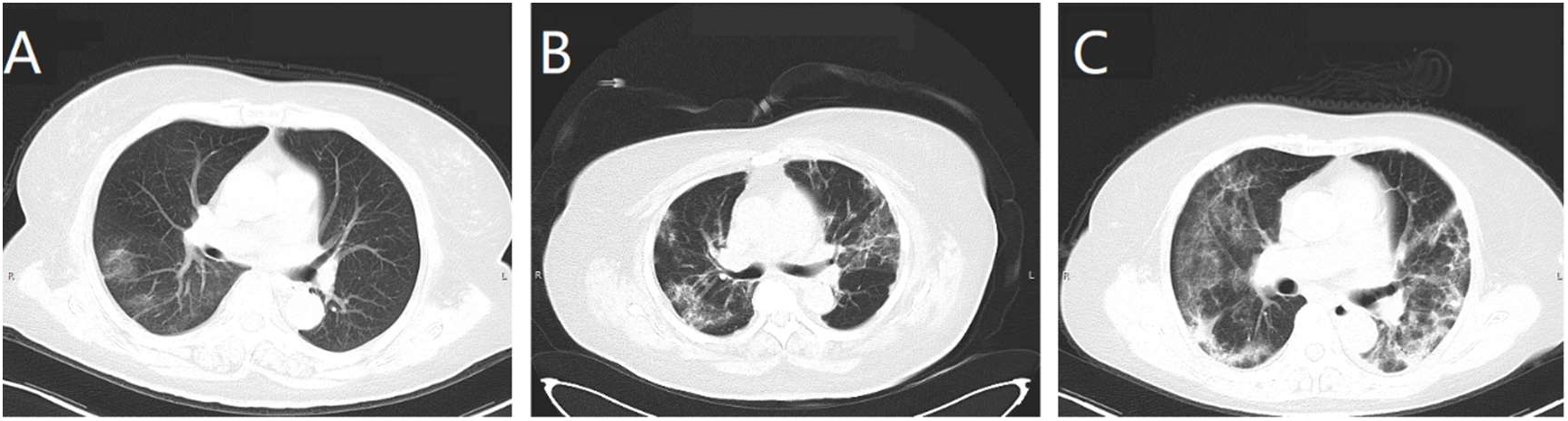
Typical CT images from each group. (A) Patient was from the mild group; this image shows local right-sided ground-glass opacity. (B) Patient was from the severe group, bilateral multiple ground-glass opacities were showed. (C) Patient was from the critical group, double-sided patchy consolidation and multiple bilateral ground-glass opacities were showed.

Most patients received antiviral and antibiotic treatment (n =33, 94·3% and n =29, 82·9% respectively), 13 (37·1%) patients received glucocorticoids (Table 2). 9 (25·7%) patients received intravenous immunoglobulin therapy. Three (8·6%) patients received anticoagulant therapy. Oxygen therapy was used for every patient. Most of the patients received oxygen by mask or nasal catheter. Only 2 (5·7%) patients received non-invasive ventilation with bilevel positive airway pressure (BiPAP), and they were free of BiPAP on the day we collected the samples. No patient received Invasive ventilation (Table 2). By the end of Feb 22, 1 (2·9%) patient had been discharged, and all other patients were still in the hospital (Table 1).

The infection rate for the 35 female patents’ sexual partner was up to 42·9%, but only 2 patients, Patient 14 and 19, admitted having sexual behavior with their partners from 14 days before the onset of first symptoms to the day we collected samples (Table 1, Table 4, Figure 2). The detailed timelines are exhibited (Figure 2). Both patients use intrauterine device for contraception. Patient 14 was 48 years old. She had regular sex behavior during the 14 days before first symptom (Figure 2). While stopped since she suffered from fever with a highest temperature above 39·0℃. Patient 19 was 41 years old. Her duration of sex behavior was longer than patient 14, which ended at the day she was diagnosed by throat swab. Her first symptom was also fever, but the highest temperature (38·9℃) is lower than patient 14 and her symptoms were mild in need of lower flow of oxygen (Figure 2). Patient 19’s partner was clinical diagnosed with COVID-19, while patient 14’s was not.

**Table 4:** SARS-CoV-2 Test by RT-PCR for Exfoliated cells samples from cervix or vaginal residue, Vaginal discharge, anal swab samples. Recent sex behavior and sexual partner infection of every patient.

| Patient NO. | Exfoliated cell samples from cervix or vaginal residue* | Vaginal discharge specimens* | Anal swab samples* | Throat swab samples* | Sex behavior† | Sexual partner infection‡ |
| --- | --- | --- | --- | --- | --- | --- |
| 1 | - | - | - | + | - | + |
| 2 | - | - | - | + | - | - |
| 3 | - | - | - | + | - | - |
| 4 | - | - | - | + | - | + |
| 5 | - | - | - | - | - | - |
| 6 | - | - | - | + | - | + |
| 7 | - | - | - | + | - | - |
| 8 | - | - | - | + | - | - |
| 9 | - | - | - | + | - | - |
| 10 | - | - | - | + | - | - |
| 11 | - | - | - | + | - | - |
| 12 | - | - | - | - | - | - |
| 13 | - | - | - | + | - | - |
| 14 | - | - | - | + | + | - |
| 15 | - | - | - | - | - | - |
| 16 | - | - | - | + | - | - |
| 17 | - | - | - | - | - | - |
| 18 | - | - | - | + | - | - |
| 19 | - | - | - | + | + | + |
| 20 | - | - | - | + | - | + |
| 21 | - | - | - | - | - | + |
| 22 | - | - | - | - | - | + |
| 23 | - | - | - | - | - | - |
| 24 | - | - | - | - | - | - |
| 25 | - | - | - | + | - | - |
| 26 | - | - | - | + | - | + |
| 27 | - | - | - | + | - | + |
| 28 | - | - | - | + | - | + |
| 29 | - | - | - | + | - | + |
| 30 | - | - | + | + | - | + |
| 31 | - | - | - | + | - | + |
| 32 | - | - | - | + | - | + |
| 33 | - | - | - | + | - | - |
| 34 | - | - | - | + | - | - |
| 35 | - | - | - | + | - | + |

\* “+” stands for positive results, “-” stands for negative results.
†Time window: from 14 days before first symptoms to the day we collected samples. “+” stands for had sex behavior recently, “-” stands for didn’t have sex behavior recently.
‡ “+” stands for infected, “-” stands for not infected.

**Figure 2:**
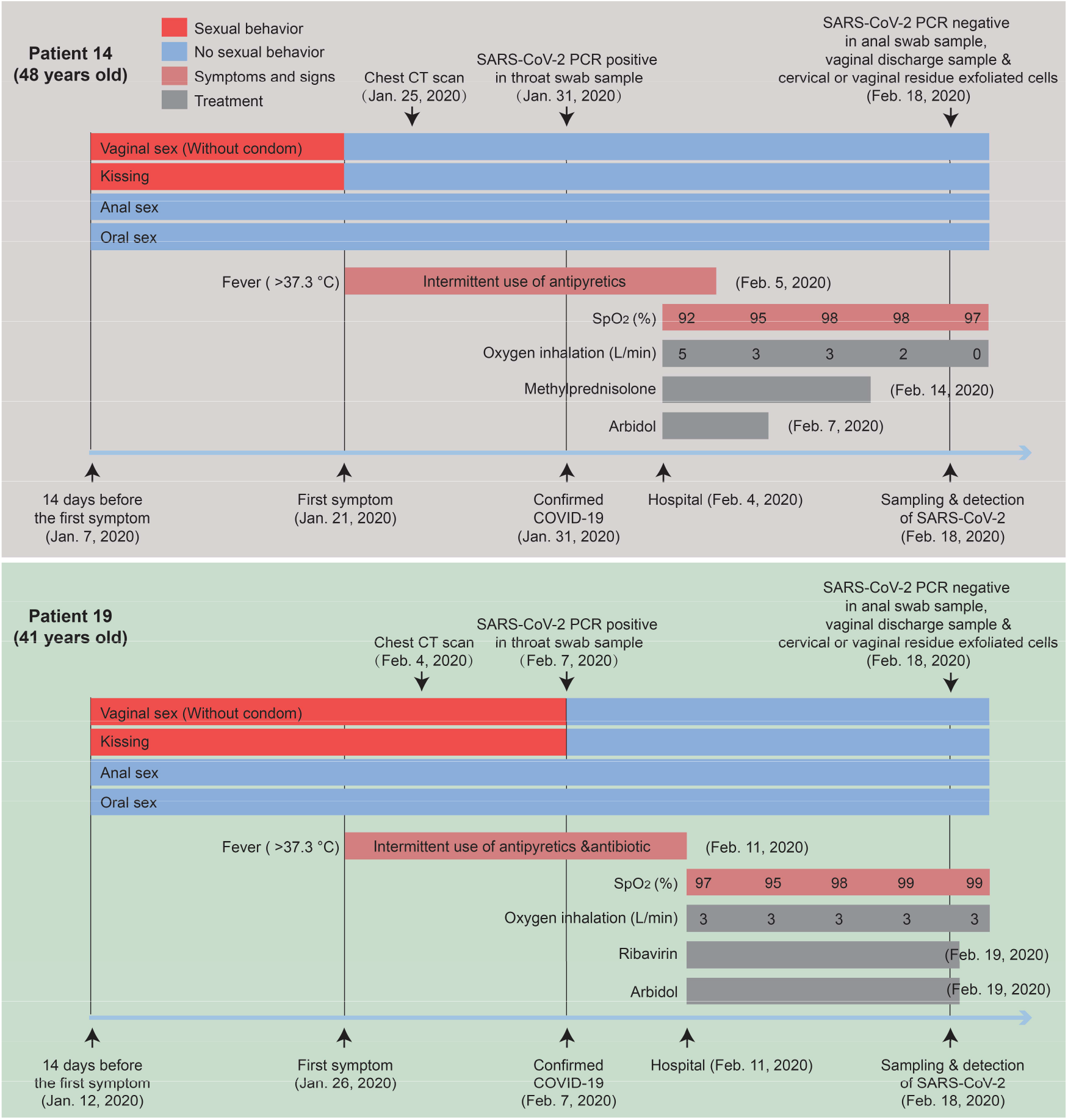
Sexual behavior history, diagnosis and treatment process of patient 14 and patient 19 with COVID-19.

Analysis of SARS-CoV-2 was conducted using the vaginal environment and anal swab samples of 35 female patients with COVID-19. All the vaginal environment samples (35 cases in total) were negative. One anal swab sample (35 cases in total) was positive (Table 4), which was from a 64-year-old female of Wuhan, who had entered menopause for 10 years. Her husband was also diagnosed with COVID-19, while they had no sexual behavior from 14 days before the first of symptoms to the day samples collected.

## DISCUSSION

The evidence so far suggests that respiratory droplets and intimate contact are the two main transmission routes of SARS-CoV-2^19-22^. Aerosol transmission also exists in relatively closed unventilated space^23^. Fecal-oral transmission is possible^23^. Vertical transmission have not been verified^16,23^, although scientists have found that SARS-CoV-2 is positive in feces and blood^15^, while negative in amniotic fluid, cord blood, neonatal throat swab, and breastmilk^16^. In addition, asymptomatic infected persons are potential sources of infection, with the long incubation period, increasing the difficulty in epidemic prevention and control^24^. Therefore, the transmission of SARS-CoV-2 is still worth exploring.

Herein, we reported a cohort of 35 female patients, with an age range of 37-88. Additionally, over 54·3% (19/35) patients infected with SARS-CoV-2 had chronic diseases including cancer (Table 1), are more susceptible due to the weaker immune function^25^, in accordance with SARS-CoV and MERS-CoV^5^. There was a special patient undergoing caesarean section due to scar uterus contractions. We obtained the vaginal secretions on the third day after operation and found the SARS-CoV-2 negative similar to breastmilk, anal swab and neonatal throat swab samples. Up to now, no evidence suggested that pregnant women need caesarean section merely because of SARS-CoV-2 positive in mother’s throat swab^16^.

Our study showed that female patients with COVID-19 have a similar trait of clinical characteristics to the patients reported recently. Most common first symptom was fever (25, 71·4%). Lymphopenia was a common phenomenon, suggesting that SARS-CoV-2 might primarily target lymphocytes, like SARS-CoV^5^. IL-2R, IL6, and TNF-α were above the normal range in half patients. A cytokine storm was induced after SARS-CoV-2 infection, followed by large amounts of immune responses and changes in immune cells such as lymphocytes, which was same as people infected SARS-CoV^5,26^. Oxygen therapy was used for every patient to improve hypoxia. Most patients received antiviral treatment and antibiotic treatment to inhibit virus replication and prevent bacterial co-infection. Glucocorticoids and immunoglobulin therapy used in some patients is in order to inhibit inflammatory response based on “cytokine storm” theory. However, some researchers suggested that the use of antibiotics and glucocorticoids are not good for patients due to lacking strong evidence^21^.

Sex behavior is a cluster of intimate contacts involving hugs, kisses, and oral/anal sex, et al. Researchers have found SARS-CoV-2 positive in throat swab, droplet, anal swab, saliva, urine, and tear samples^13,23,27^, whether female vaginal environment contains SARS-CoV-2 triggered our interest. We collected 105 samples from 35 patients to detect the SARS-CoV-2, including vaginal discharge, cervical or vaginal residue exfoliated cells and anal swap, It is worth noting that only an anal swab sample from one patient was positive for SARS-CoV-2, which is in consistence with the possibility of fecal-oral transmission reported previously ^23^. The unexpected results from the vaginal environment may due to the negative expression of ACE2, the receptor of SARS-Cov-2, in the vagina and cervix^28^.

Additionally, for two female patients admitted having sexual behaviors in the possible incubation period with their partners, one patient’s partner was diagnosed with COVID-19. We cannot conclude that the infected partner is transmitted by her through sex behavior, as the samples of her vaginal environment anal swab samples were all SARS-CoV-2 free. But we should consider sex behavior as a cluster of intimate contacts involving hugs, kisses, and oral/anal sex, et al. She might transfer the virus to him by these intimate contacts, or the other way around. It is worth noting the infection rate for the 35 female patents’ sexual partner was up to 42·9%, which is not a low proportion. Given to the fact that females are shy to talk about sex in China, it was probably many patients had hidden their sex history. So, sex partners got infected by the intimate contacts during sex might be more than one in the 42·9% group. If one of the involvers in a sex behavior is in incubation period or is asymptomatic patient, the risk of the other involver got infected is pretty high. As asymptomatic persons are potential sources of SARS-CoV-2 infection^24^.

Transmission between family members occurred in 13-21% of MERS cases and 22-39% of SARS cases^5^. Moreover, many patients with SARS or MERS were infected through super-spreaders ^5^. Investigators had reported that the main route of SARS-CoV transmission was via respiratory droplets, and may also be transmitted through fecal-oral and contiguous pathways^23^. Unlike SARS-CoV, the main transmission route of MERS-CoV was large droplets and contact. It is still not clear whether SARS-CoV or MERS-CoV is transmitted through sexual transmission. Based on the transmission route of SARS-CoV and MERS-CoV, preventive measures, including maintaining good personal and environmental health, and implementing strict contact and droplet prevention measures in the community, can effectively prevent the spread of SARS-CoV and MERS-CoV. In this study, no evidence indicated that SARS-CoV-2 could transmit by vaginal sex from female to her partner. But the risk of infection by oral/anal sex or other intimate contacts during sex should not be ignored. We suggest that people in the affected areas should be very cautious about sex behavior, avoid oral/anal sex especially. It might be helpful to reduce the global spreading of SARS-CoV-2.

Our study has several limitations. First, the sample size was relatively small in which only 35 patients with SARS-CoV-2 positive or CT suspected was included; much more patients including sexually active women are needed to confirm our findings in the future. Second, samples from partner of the patients enrolled is missing, including anal swabs, semen, and urethral orifice swabs. Nevertheless, the data in our study revealed the epidemiological and clinical characteristics of female patients with COVID-19 in a new prospective. In summary, though no evidence was found in this study that SARS-CoV-2 can be transmitted by vaginal sex from female to the partner. Considering the complexity of sexual behavior and the transmission potential of asymptomatic infectives, we suggest that people who lived in the epidemic areas, have tourism history from epidemic areas and were potentially asymptomatic infected should be very cautious about sex behavior and avoid oral/anal sex especially, which may be helpful to prevent further spreading of SARS-CoV-2.

## Data Availability

With the permission of the corresponding authors, we can provide participant data without names and identifiers, but not the study protocol, statistical analysis plan, or informed consent form. Data can be provided after the article is published. Once the data can be made public, the research team will provide an email address for communication. The corresponding authors have the right to decide whether to share the data or not based on the research objectives and plan provided.

## Contributors

PC, X Ma, and SW made substantial contributions to the study concept and design. PC, ZC, TW and JD took responsibility for obtaining written consent from patients, obtaining ethical approval, collecting samples, and confirming data accuracy. CZ, WS, SW and TD participated in data collection. JL was in charge of the laboratory tasks, including sample processing and detection. PC, ZC, TW and JD contributed equally and share first authorship. X Ma and SW contributed equally to this article had full access to all of the data in the study and take responsibility for the integrity of the data and the accuracy of the data analysis. PC, ZC, TW and JD participated in drafting the manuscript, and revising it on the basis of reviewers’ comments. ZC, JD and JJ made substantial contributions to data acquisition, analysis, and interpretation. SW, X Ma, JC, YR and X Miao revised the final manuscript.

## Acknowledgments

This study was supported by the Clinical Research Pilot Project of Tongji hospital, Huazhong University of Science and Technology (No. 2019CR205). We thank all patients involved in the study.

